# Detection of Malaria Infection from parasite-free blood smears

**DOI:** 10.64898/2025.12.29.25343125

**Authors:** Nicolas Bourriez, Saranga Kingkor Mahanta, Ivan Svatko, Eleanor Lacassagne, Aurore Atchade, Francesco Leonardi, Ethan Cohen, Achille Massougbodji, Nicolas Argy, Gilles Cottrell, Auguste Genovesio

## Abstract

Malaria affects almost 263 million people worldwide, most of whom live in sub-Saharan countries. In a strategy to reduce malaria-related mortality and limit transmission, diagnosis in endemic areas needs to be immediately available on the field, easy to perform and cheap. Therefore, it currently heavily relies on microscopic examination of blood smears. However, several studies comparing the sensitivity of this approach with qPCR, considered as the most sensitive method albeit not available on the field, found that up to half of the infected population failed to be detected by microscopy alone because no visible parasites could be found in blood smears. These so-called *submicroscopic* infections pose a diagnostic challenge, yet represent a huge reservoir for malaria transmission.

In this study, we hypothesized that qPCR results could be predicted by deep learning from subtle cell signals present in thin blood smear images, even in the absence of visible parasites, making a sensitive diagnostic directly available on the field using a microscope and a smartphone. To test this hypothesis, we acquired a large smartphone-based blood smear images dataset from samples tested both for microscopy and qPCR. We then focused exclusively on these “negative” slides from the microscopic diagnostic point of view, among which half were qPCR positive. A range of standard deep learning models were evaluated to best predict the qPCR result from these microscopy images, using various backbones along with various aggregation functions at the slide level, from a simple vote to Multiple Instance Learning with attention.

Our results show that the qPCR results can be predicted from parasite free blood smear images with 62.00% (±2.5 on 4-folds) accuracy and reaching 67.2 % (±9.6 on 4-folds) in sensitivity. We then used generative models to investigate the subtle morphological variations occurring in red blood cells that may contribute to predicting malaria infection in the absence of parasites. Leveraging thin blood smear and portable deep learning, we established the first proof of concept that the qPCR sensitivity can be approached through the detection of submicroscopic infections directly on the field without additional infrastructure and thus could significantly improve malaria surveillance and elimination efforts.

## Introduction

Sensitive diagnosis of malaria infections deployable on the field remains a cornerstone of global malaria control and elimination initiatives. Advances in molecular diagnostics, particularly qPCR and loop-mediated isothermal amplification (LAMP) offer sensitive detection methods (Okell et al. 2012; Mosha et al. 2013; Wu et al. 2015; Notomi et al. 2000; van Eijk et al. 2023). However, their reliance on specialized equipment, higher costs, and longer processing times limits their applicability in resource-constrained field settings. While HRP2-based Rapid Diagnostic Tests (RDTs) achieve good sensitivities for moderate to high parasite densities (Abba et al. 2011), their performance deteriorates considerably at low parasitemia. In addition, progress against malaria is now threatened by an increase in *pfhrp2* and/or *pfhrp3* gene deletions resulting in false-negative RDT results (Watson et al. 2025). Consequently, microscopic examination of stained blood smears remains the gold standard diagnostic method, offering compelling advantages: low-cost, rapid results, requiring minimal infrastructure while being performed directly on the field nearby patients (Ashraf et al. 2012; Hung et al. 2017; Mathison and Pritt 2017). When parasites are visible, microscopic blood smear examination enables reliable diagnosis through well-established criteria (World Health Organization 2010; Garcia 2010). However, this approach also faces a fundamental limitation: a significant proportion of active infections are *submicroscopic*: no parasites are visible under microscopy despite their trace being detectable by qPCR, the most sensitive molecular methods (Okell et al. 2012; Mosha et al. 2013; Wu et al. 2015; Notomi et al. 2000). A recent study of over 900 individuals in South Benin highlighted this gap: qPCR revealed a prevalence of 53.8%, whereas microscopy detected only 23.4% (Telfils et al. 2024). More than half (55.6%) of positive cases were submicroscopic infections misclassified as negative by microscopy examination. This large sensitivity gap between the gold standard microscopy approach available on the field and laboratory-based qPCR, represents a significant challenge for malaria control, as these submicroscopic infections highly contribute to disease transmission (Lin Ouédraogo et al. 2015). There is therefore an urgent need for a malaria diagnostic tool that combines the high sensitivity of molecular methods with the portability and low cost of microscopy on the field. The search for such a diagnosis tool has thus become a priority of the WHO in the global fight against malaria.

Portable computer vision solutions for malaria diagnosis from blood smear images have emerged, but so far exclusively rely on parasite detection, thus only performing faster the microscopy test currently available on the field, not better. Early work by (Rajaraman et al. 2018) showed that pretrained CNNs could match expert microscopists in detecting visible parasites. Other studies by (Das et al. 2013; Yang et al. 2020) have successfully integrated these parasite detection algorithms into smartphone-based diagnostic tools, opening an interesting path towards a portable diagnosis tool in the field. While these early methods achieved a good accuracy at detecting parasites in controlled settings, they often struggled with real-world variability in sample preparation and imaging conditions (Rajaraman et al. 2018; Yang et al. 2020). More recent work has explored domain adaptation techniques to address these challenges, attempting to create models robust to variations in staining protocols and microscope configurations using optimal transport (Lyu et al. 2021). Despite these advances in architectural sophistication, these approaches remain fundamentally limited by their focus on detecting human-annotated visible parasites. By mimicking human observation, they may help fasten the microscopist work, but still inherently miss the entire submicroscopic reservoir (Lyu et al. 2021; Torres et al. 2018).

Deep learning excels at capturing subtle visual patterns that might be imperceptible to human observers. When phenotypic changes are too subtle to be discernible by humans or discriminative features cannot be reliably segmented, deep learning models can still be trained directly on full field-of-view images and achieve high accuracy (Scheeder et al. 2018; Ando et al. 2017). Furthermore, self-supervised learning models like DINOv3 (Siméoni et al. 2025), CLIP (Radford et al. 2021), and MAE (He et al. 2021) have demonstrated remarkable capabilities in learning rich visual representations without explicit labels, that can be effectively transferred with or without fine tuning to medical imaging (Raghu et al. 2019; Kim et al. 2022; Litjens et al. 2017; Fillioux et al. 2023). It allows pre-trained models to adapt to specific diagnostic tasks while maintaining robustness across different imaging conditions (Esteva et al. 2017; McKinney et al. 2020). Such an approach is particularly relevant for field-deployable solutions where image quality and acquisition conditions may vary significantly. While these approaches have transformed computer vision tasks across many domains (Yosinski et al. 2014), their application to detecting invisible biological phenotypes in infectious diseases remains limited (Moen et al. 2019; Bourriez et al. 2023; Lamiable et al. 2023).

In this work, we demonstrate that microscopy images of thin blood smear slides contain *submicroscopic* malaria infection signals. To achieve these results, we built a large dataset of smartphone-acquired parasite-free thin blood smear images with their associated qPCR positive or negative results. By systematically investigating the performance of multiple deep learning architectures at predicting qPCR results from parasite free microscopy images, we demonstrate that thin blood smears contain *submicroscopic* malaria infection signals. Finally we identify cell image features whose distribution is modified by subtle cell morphological variations and a way to visualize them using generative models. These insights confirm that submicroscopic infection induces measurable but imperceptible phenotypic change in red blood cells, yet detectable by deep learning models. Furthermore, because our proof of concept is not based on sophisticated scanner based image acquisition but on smartphone-based optical microscopy images easily available and usable on the field, we anticipate that it could be exploited for the deployment of a rapid, cost-effective diagnostics tool in resource-limited settings, where it is most needed.

## Results

### A large parasite-free microscopy image dataset from the field

To our knowledge, there is no available dataset of thin blood smear microscopy images that would be assessed as parasite free, thus specifically targeting the distinction between *negative* and *submicroscopic* cases. Therefore, establishing a quantitative proof that *submicroscopic* cases could be detected from parasite-free microscopy images was so far impossible. To address this question, we organized the image acquisition of a large-scale dataset of thin blood smear samples collected through a field study on asymptomatic (fever free) participants in Benin (Telfils et al. 2024). Each slide derived from a study participant and was categorized on the field by a human operator as either microscopy positive (visible parasites), submicroscopic (qPCR-positive but microscopy-negative), or negative (qPCR-negative, and hence microscopy negative). To focus our experiment on the ability of a deep learning model to detect infection in the absence of visible parasites, we considered parasite-free slides only (**Figure 1**a). We then performed qPCR test and smartphone based image acquisition of thin blood smear from these slides (**Figure 1**b), ending up in 10 Fields-of-View (FoVs) from each 316 submicroscopic and 316 negative slides for a total of 6320 FoVs (**Figure 1**c) with a slide level qPCR annotation (**Figure 1**d). Image capture was performed using a smartphone mounted on a Leica DME microscope at ×1000 oil immersion magnification available on the field. To ensure consistency in image quality, a small convolutional neural network (CNN) was embedded within a custom application in the smartphone to automatically assess on the fly the slide quality prior to final acquisition. Examples of image are provided in **Supplementary Fig.1**.

**Figure 1:**
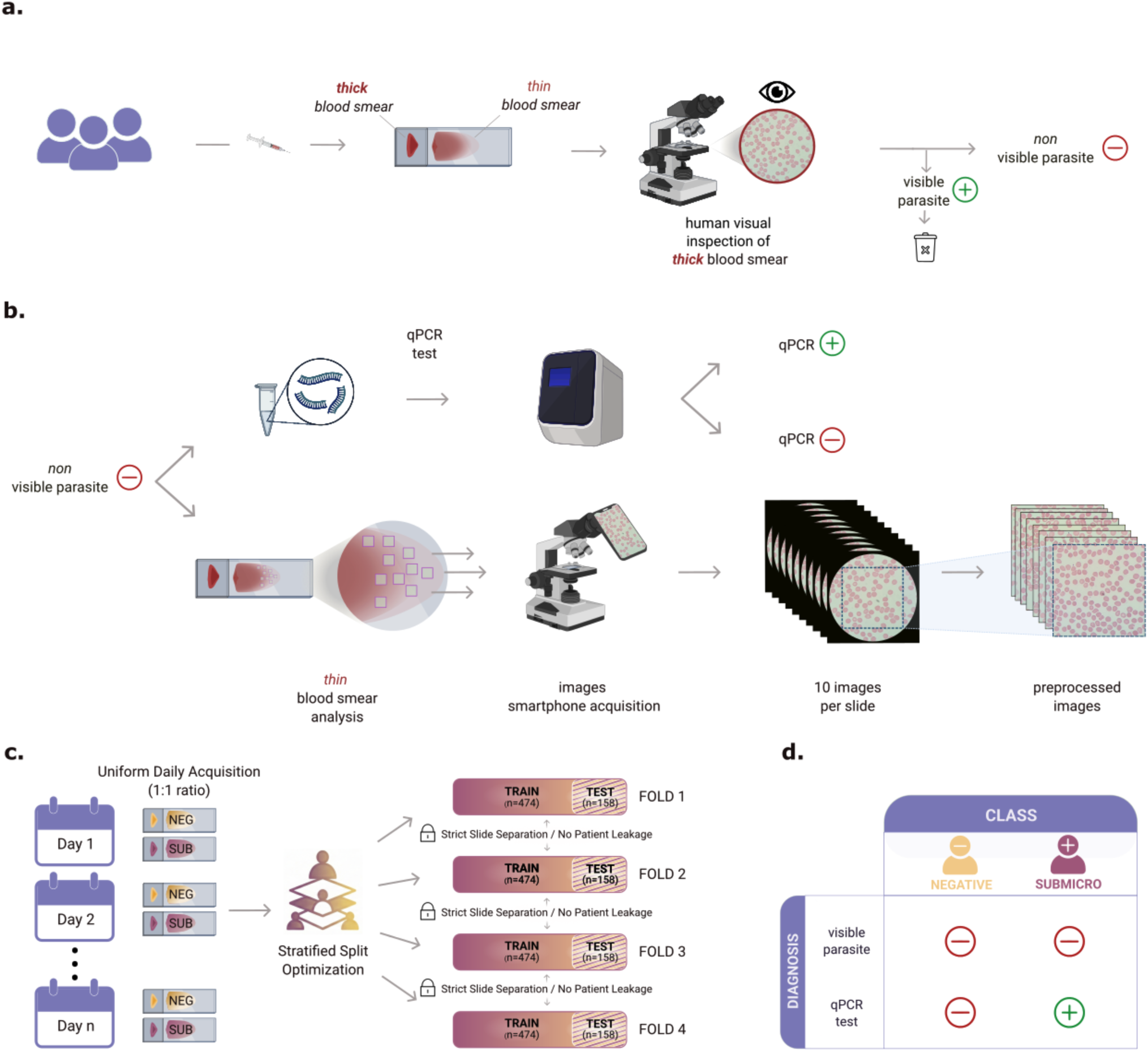
Overview of samples collection, diagnostic workflow, and images acquisition for submicroscopic malaria detection using deep learning. **a.** Blood samples were collected from asymptomatic participants and a first diagnosis was performed on the field through thick blood smear visual inspection by a microscopist. Samples without any visible parasite were selected to build our dataset. **b.** A quantitative PCR (qPCR) test was performed off the field and thin blood smear images were subsequently acquired using a smartphone mounted on a Leica DME microscope at x1000 oil immersion magnification (10 Fields-of-View per slide) **c.** To mitigate experimental batch effects, image acquisition followed a strict daily paired protocol (equal number of Negative and Submicroscopic slides per day). An equal number of 316 slides per class were acquired. The resulting dataset was partitioned into four folds using an optimization algorithm designed to minimize the average quadratic deviation from a perfect daily class balance ($ratio=0.5$). Each fold comprises 474 training and 158 testing images. Splits were stratified by Slide ID to ensure strict patient independence and prevent data leakage between training and testing sets. **d.** Class affectation was exclusively based on qPCR results since all slides were considered negative by the microscopist on the field.

### Infection detection from parasite-free thin blood smear images

Equipped with this new dataset, we could investigate if a deep learning model could predict the submicroscopic and negative qPCR results from microscopy images of thin blood smears. Our goal here was not to develop an optimal deep learning architecture but rather to establish a proof of concept that a submicroscopic signal is detectable in the absence of visible parasites. We then tested a range of encoders for the individual FoVs, from simple CNN trained from scratch to frozen foundation models such as DINOv3. We then passed these encodings through a simple MLP to predict the individual FoV probability for these two classes (Negative vs. Submicroscopic). We aggregated the 10 FoVs’ probabilities per slide to a slide-level prediction, evaluating various methods from voting to attention-based aggregation (**Figure 2**a). All models and aggregation combinations were compared based on a 4-fold cross validation procedure (**Figure 2**b). We provide details of the results per fold (**Supplementary Fig.2**) along with the ROC curves (**Supplementary Fig.3**). The combination with the best accuracy reached 62.0% (±2.5 on 4-folds) with a specificity of 56.7% (±07.0) and a sensitivity of 67,2% (±9.6), relative to qPCR. Interestingly, most models beyond custom standard CNN, could detect the submicroscopic signal as attested by their non-random accuracy, suggesting that such a model can capture subtle phenotypic signals from parasite-free slides (**Supplementary Table 1**). We further investigated if a lower number of FoVs could be used while maintaining similar diagnosis performances. Compared to 62.0%, the accuracy of the best model, we show that the same model (trained on 10 FoVs per slide) maintains an accuracy above 60.0% with 5 FoV per slide, demonstrating that the signal is probably diffused (**Figure 2**c). We provide details per fold (**Supplementary Fig.4**). Still, increasing the number of FoVs slightly improves the results and performance stabilizes beyond 8 FoVs per slide, with no substantial gains with additional FoVs. These results establish for the first time the existence of a detectable image-based signature associated with submicroscopic malaria infection phenotypes that can be leveraged in the absence of visible parasites.

**Figure 2:**
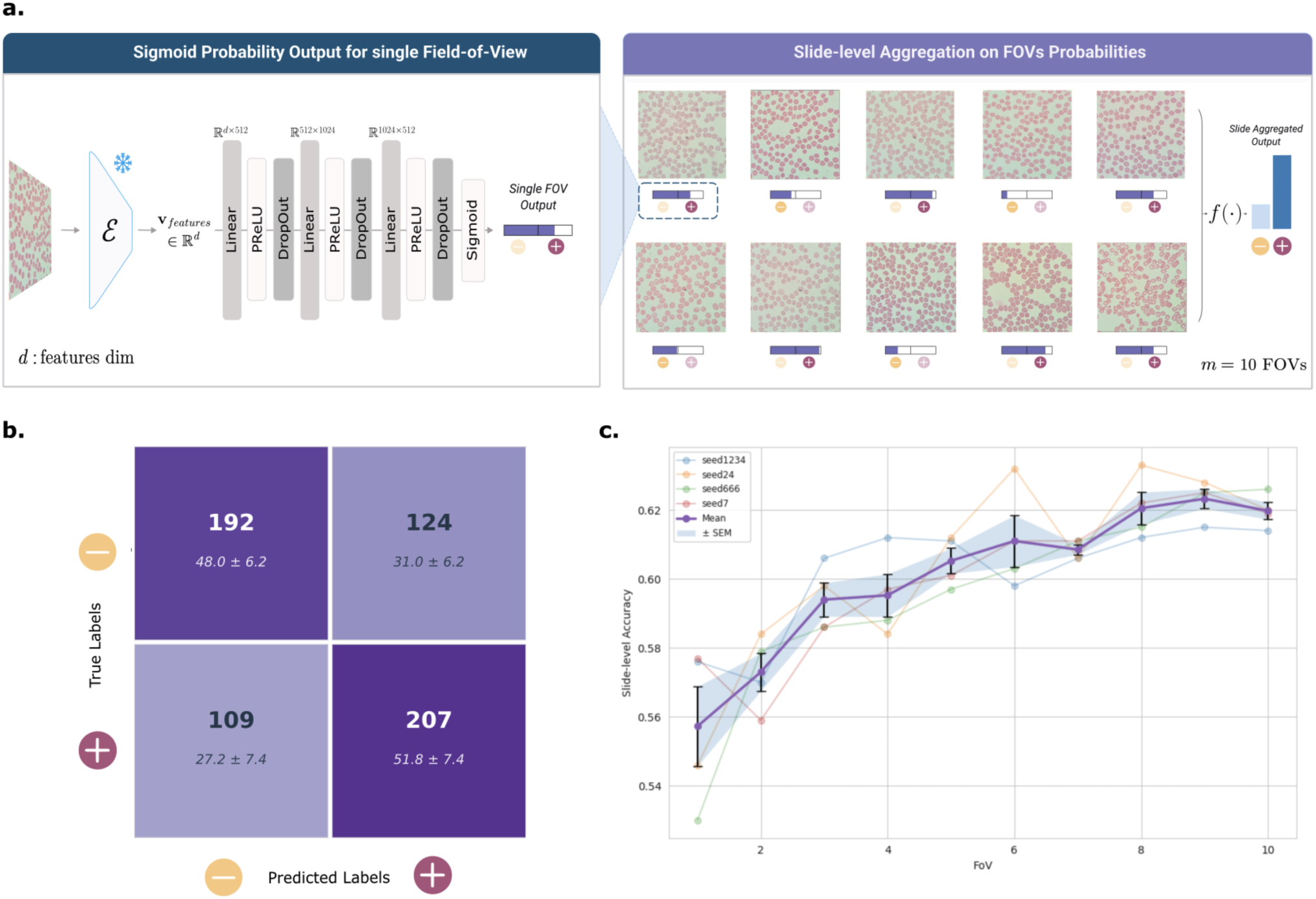
Deep learning enables reliable detection of submicroscopic malaria infections from thin blood smear images. **a.** Schematic of the classification pipeline: each FoV of thin blood smear is processed through a feature extractor to generate a class token embedding, which is passed through a fully connected layer to produce a sigmoid output for the binary classification (Negative vs. Submicroscopic). The predictions from the *m* Fields-of-View of a slide are then aggregated to obtain a final slide-level classification. Various encoder architectures and aggregation methods could systematically detect *submicroscopic* infection **b.** Confusion matrix of the best-performing model summed over the 4-folds test sets, illustrating classification performance across *negative* (NEG) and *submicroscopic* (SUB) cases. **c.** Best performing model accuracy on the 4-folds & 4 seeds as a function of the number of FoVs used at inference.

### Deciphering invisible submicroscopic phenotypes

The previous classification results demonstrated that a detectable signal difference is present in images of thin blood smear to distinguish submicroscopic from negative slides. However, these subtle variations in cell phenotypes are imperceptible to human observers. We first performed a segmentation of all cells from the dataset and computed 106 quantitative features from these segmentations (FormFactor, lower Eccentricity, higher Solidity, compactness and central pallor etc.). We then performed systematic KS-tests to identify what feature had its distribution modified by the infection (**Figure 3**a). We thus uncover that many of these feature distributions were slightly but statistically significantly affected by the infection (**Supplementary Fig.5** and **Supplementary Fig.6**). In order to illustrate the effect of this subtle subpopulation shift, we train a conditional generative model using the class label as condition (Lamiable et al. 2023; Bourou et al. 2023). We then generated a large amount of paired cell images using the two conditions on the same random seed. We then applied the same cell segmentation and measurement pipeline to select generated cell pairs which displayed the largest shift on the features that had the largest KS statistics (**Figure 3**b). Although these effects are subtle shifts in subpopulations, pointing at these extreme examples helped illustrating visually the morphological effects captured by each of these features on our own data (**Figure 3**c). We could then group these features into two main categories: some of them reported a slight increase in the roundness of infected red blood cells while the other tended to point to an increase of the central pallor of the cell. These results provide a potential mechanistic explanation for the classifier’s performance. The increased central pallor observation is also supported by findings in (Chaves et al. 2025), who demonstrated in the same study that asymptomatic malaria infections — including submicroscopic cases — were associated to a lower hemoglobin levels and an increased risk of anemia, particularly among young children.

**Figure 3:**
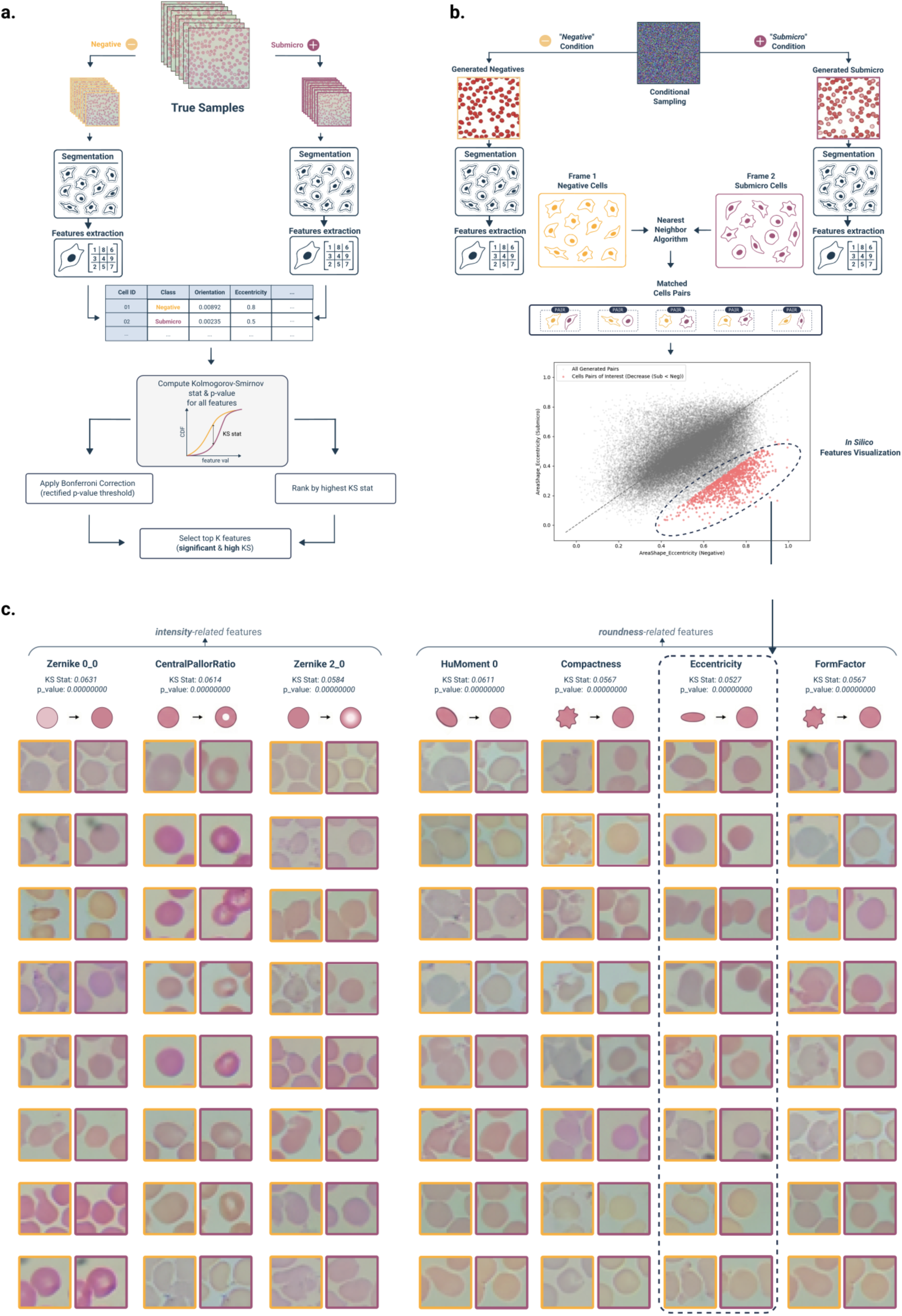
Generative discovery and statistical validation of submicroscopic malaria phenotypes. **a.** To characterize the submicroscopic signal, morphological features were extracted from real Negative and Submicroscopic cells. The robustness of candidate biomarkers (e.g. FormFactor, Eccentricity) was assessed using a threshold sweep, showing that the Odds Ratio (blue) remains > 1 and the p-value (red) decreases significantly. A FoV-level permutation test confirms the signal is not caused by batch effect if the observed difference in outlier frequency (red line) falls well outside the null distribution generated by random label permutation (p<0.001). **b.** A conditional diffusion model was used to generate counterfactual “twin” FoVs (Negative vs. Submicroscopic) from shared noise distribution. Comparison of segmented cell pairs in the synthetic domain reveals that the model induces specific morphological changes to translate from *negative* to *submicro* class, such as increased *roundness*, *compactness* and *central pallor.* **c.** Synthetic counterfactual pairs display the predicted morphological shift of *lower eccentricity*.

## Discussion

In this study, we built a large thin blood smear image dataset acquired in realistic field conditions that includes negative and submicroscopic (qPCR positive) cases. By rigorously evaluating various deep learning architectures on 4-fold cross validation, we demonstrated that thin blood smear images contain latent signals that, even in the complete absence of visible parasites, can be leveraged to distinguish submicroscopic infections from negative samples. The best of these configurations reached 62.00% (±2.5 on 4-folds) accuracy with a specificity of 56.7.0% (± 7.0) and a sensitivity of 67.2 % (±9.6 on 4-folds) compared to qPCR which is very encouraging when compared to microscopy alone that reach only 43.5% (Telfils et al. 2024). We further investigated the variations in cell phenotypes triggered by the infection and identified a set of quantitative features showing a distribution shift likely due to a subpopulation shift.

These results indicate that submicroscopic infections induce measurable alterations in red blood cells, detectable via deep learning despite being imperceptible to human observers. This suggests a qPCR-level sensitivity could be approached directly in the field without complex infrastructure. Such a tool would be of crucial importance in terms of progress in the fight against malaria, knowing that the search for portable and more sensitive diagnostic tools is a priority recommended by the WHO to achieve the elimination of malaria.

Recent work has shown that malaria parasites actively remodel host red blood cells (RBCs) through secreted factors, altering membrane deformability and cytoskeletal organization (Dekel et al. 2021; Scovino et al. 2022). For instance, *Plasmodium falciparum* secretes functional 20S proteasome complexes within extracellular vesicles to modify the mechanical properties of host red blood cells (Dekel et al. 2021), building on previous work showing parasites actively remodel host cells through multiple molecular pathways, including phosphorylation cascades (Zuccala et al. 2016), cytoskeletal modifications (Zhang et al. 2015), and membrane protein trafficking (Sisquella et al. 2017; Suárez-Cortés et al. 2016). These studies reveal how host cell remodelling from parasites (Gilson et al. 2017; Chandramohanadas et al. 2011) alter red blood cell deformability and membrane properties even before they become detectable by microscopy. The latter suggests information exists beyond the mere presence of parasites, encoded in subtle host cell shape variations. Interestingly, other studies have even shown that in a *Plasmodium* infection environment, morphological changes affect not only infected red blood cells but also uninfected ones (Paul et al. 2013). This bystander effect could be, at least in part, responsible for the detection of submicroscopic infections from parasite-free slides. While these morphological changes are invisible to microscopic operators due to natural cell-to-cell variability(Arevalo et al. 2024), they offer a potential target for an enhanced deep learning analysis.

Notably in our study, some cases labeled as *negatives* by qPCR were classified as *submicroscopic* by the model. While it is accounted here as misclassification, it may also reflect the limit of qPCR precision at very low parasitemia levels. The heteroskedasticity of qPCR measurements, wherein precision decreases as parasite density approaches the detection limit, raises the possibility that some predicted submicroscopic cases are biologically meaningful despite falling below qPCR’s threshold. This highlights a key challenge in validating deep learning predictions against molecular reference methods, particularly when both approaches operate near the lower bounds of detectability.

While the point of this work was to present a theoretically sound proof of concept for a sensitive malaria diagnosis directly available on the field, its practical implementation for real-world use - particularly on smartphones - would require further engineering efforts including model compression and on-device inference efficiency. Beyond implementation challenges, several technical limitations must be addressed. One key issue is the generalization capabilities of the model across variations in slide staining, smartphone cameras, microscope configurations and operators, as these may introduce image distribution shifts that may affect deep learning model decision. Future work will focus on expanding the current dataset to improve generalization across diverse transmission settings by increasing the diversity of training samples, particularly from different geographic regions. We will also optimize the model architecture to enhance both accuracy and computational efficiency, to enable real-time deployment on smartphones with resource-constrained hardware through model distillation. Another important aspect of future research will be to improve the interpretability of deep learning models. While we have taken initial steps toward understanding which biological features could contribute to the model’s decision-making process using conditional image-to-image translation, further work would be required to develop even more descriptive approaches.

In conclusion, we provided the first proof of concept that submicroscopic malaria infection detection is possible via a deep learning algorithm, combined with the detection of subtle plausible phenotypes according to the literature (central pallor and morphological modification of the red blood cell membrane). We have, for the first time, broken the glass ceiling of previously developed artificial intelligence-based malaria diagnostic methods, all of which are based on searching for and counting the parasite, much like a human microscopist. Beyond malaria diagnosis, this work intends to highlight the potential of deep learning for invisible pattern analysis of blood cells. Further research could focus on refining these models for prediction of standard blood panels such as lipid, hemoglobin or glucose tests in limited resource settings.

## Methods

### Sample collection and acquisition

Blood smear slides used to acquire the image collection necessary for the development of deep machine learning were obtained from the field study by Telfils R et al. in Benin, using standard laboratory procedures for malaria diagnosis (Telfils et al. 2024). At inclusion, in parallel to RDT screening based on PfHRP2 antigen detection, whole blood sampling was performed for malaria microscopic diagnosis and parasitaemia quantification made respectively through standardized Lambaréné thick drop–blood smear (TBS) and thin blood smear using standard Giemsa staining protocols. TBS were considered positive if at least one trophozoite was visualized after examination of 1,000 leukocytes. TBS were read by 2 microscopists and any discordances between readers or between techniques (microscopy vs. qPCR) were confirmed by a third microscopic examination. To confirm or exclude malaria infection and precise involved *Plasmodium* species, *Plasmodium* DNA detection was performed using a real-time quantitative polymerase chain reaction (qPCR) targeting *P. falciparum* cytochrome b mitochondrial sequence, *P. malariae* 18S rRNA sequence and *P. ovale* dihydrofolate reductase gene as already described (Telfils R et al., 2024).

Digital images acquisition was retrospectively performed in France from this thin blood smear collection using a Leica DME microscope-mounted smartphone at high-magnification (×1000 oil immersion) to reproduce the standard practice of microscopic examination for the diagnosis of malaria infection. Ten Fields-of-View per slide were randomly captured in the well coloured and smooth end spread of the smear where the red blood cells are side by side and overlap as little as possible. Image quality check control system is embedded in the smartphone application to alert users about the low image quality (blurry or crowded images as well as staining artefacts or poor cell staining) needed to restart image acquisition to increase their usefulness in deep learning model development.

### Image preprocessing

To standardize image inputs while preserving diagnostically relevant information, we applied a contour-based preprocessing pipeline to extract the central region of interest (ROI) from each microscopy image. This process involved grayscale conversion, thresholding, contour detection, masking, and cropping, ensuring that only the relevant blood smear area was retained. To enable the use of pretrained encoders, images were normalized following standard practices. First, pixel values were rescaled to the [0,1] range, then normalized using the mean and standard deviation values of the ImageNet dataset, ensuring compatibility with models pretrained on this distribution. In addition, data augmentation - random horizontal and vertical flips - was applied to improve model generalization.

### Feature extraction

For feature extraction, we employed transfer learning, freezing the pretrained backbone and training only the classifier head. The custom CNN was the only architecture trained from scratch, and results (**Supplementary Table 1**) confirm that training from scratch significantly underperforms compared to transfer learning approaches. While alternative fine-tuning strategies such as full fine-tuning, gradual unfreezing, and LoRA were explored, they did not yield notable improvements or resulted in overfitting and are thus omitted from the main results. To compare architectures with fundamentally different designs, we used ResNet50, a residual convolutional neural network (CNN), and DINOv3, a self-supervised vision transformer (ViT). Unlike CNNs that operate through hierarchical spatial filters, ViTs process images as sequences of patches and rely on global self-attention mechanisms to capture long-range dependencies. ResNet50 was pretrained in a supervised fashion on the ImageNet dataset (∼1.3M images), limiting its exposure to visual variability. In contrast, DINOv3 was pretrained on a large-scale corpus of 1.7 billion images in a self-supervised manner, offering rich and generalizable representations. To ensure a fair comparison across encoders, all models were trained with identical hyperparameters (optimizer, learning rate, weight decay, batch size) and used the same classifier head.

### Slide-level aggregation and classification

Thin blood smears slides were analyzed at the Field-of-View (FoV) level, with each FoV processed independently through the feature extraction backbone. Given a set of extracted patch features 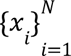, where *N* denotes the number of sampled patches from a slide, we explored different aggregation strategies to derive a slide-level prediction.

In the **voting-mechanism approach,** predictions from individual Fields-of-View (FoVs) contributed to the final slide diagnosis based on a majority vote. Given a set of *m* extracted FoVs from a slide, each indexed by *i*, the model produced a probability distribution over the *C* possible classes for each FoV, denoted as:

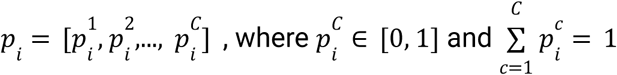

For each FoV, the predicted class was determined as the argmax of the probability distribution:

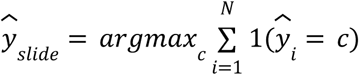

where 1(·) is the indicator function that counts occurrences of class *c* among the FoV-level predictions, *N* is the total number of FoVs and 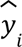 is the predicted class for the *i^th^* FoV.

In the **mean-probability approach**, the final slide-level prediction is obtained by averaging the probability distributions over all available FoVs. Given a set of *m* FoVs extracted from a slide, where each FoV *i* produces a probability vector 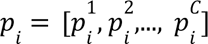 over the *C* possible classes, the aggregated slide-level probability is computed as:

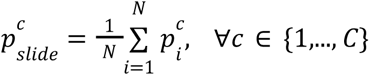

The final classification decision is then determined as the class corresponding to the maximum probability:

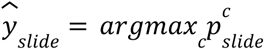

In the **median-probability approach**, the same formula is applied but the median per class is taken rather than the mean.

In the **deepsets approach,** introduced by Zaheer et al. (Zaheer et al. 2017), a permutation-invariant neural network is designed for set-based learning. Given *N* patch-level embeddings *h_i_* of dimension *D*, the DeepSets aggregation is defined as:

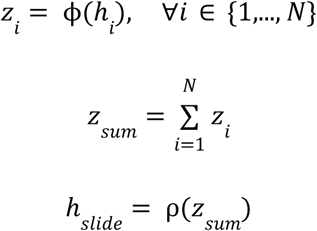

where ϕ: *R^D^* → *R^D^* is a learnable nonlinear transformation, and ρ:*R^d^* → *R^D^* is another learned transformation. Since summation is permutation-invariant, this method ensures that the final representation does not depend on the ordering of the patches.

Another permutation-invariant slide-level learning approach used is **Top-K pooling** on patch embeddings based on their L2 norm. It selects the *k* most “important” patches (those with the largest embedding norm) and then averages them into a single feature vector, a global descriptor of the slide.

Given a slide represented by *N*_*i*_ patch embeddings (*h*) obtained by passing *N* FoVs (corresponding to a slide), the importance of each *h_i_* is quantified by the L2 norm of its embedding:

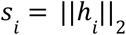

The indices of the *k* patches with the largest norms are then selected:

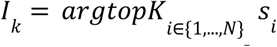

The selected Top-*k* patch embeddings are aggregated by averaging to obtain a slide-level representation:

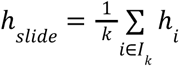

In the **gated attention approach,** introduced by Ilse et al. (Zaheer et al. 2017; Ilse et al. 2018) as a Multiple Instance Learning framework, an attention mechanism is designed to assign different attention weights to patches. Unlike averaging, attention is built to focus on the most relevant patches. Each patch embedding *h_i_* undergoes a gating mechanism using two distinct transformations:

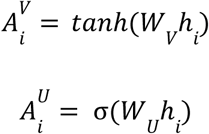

where *W_V_*, *W_U_* ∈ *R*^*D*×*D*^ are learnable weight matrices. The *tanh* function introduces nonlinearity, while σ gates the information flow. Following these two transformed representations, we then compute the attention scores as:

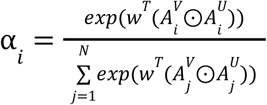

where *w* ∈ *R^D^* is a learnable weight vector and ⊙ denotes element-wise multiplication. We compute the final slide-level representation as a weighted sum:

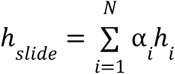

In the **transformer attention approach** (without positional embeddings), inspired by the classical Transformer architecture (Vaswani et al. 2017), we apply self-attention across patches to learn their relationships. Unlike the standard design, positional embeddings are omitted to force the permutation invariance.

We introduce a learnable class token (*z_CLS_* ∈ *R^D^*), randomly initialized, that serves as the global representation of the slide. The patch embeddings *X* ∈ *R*^*N*×*D*^(where *N* is the number of patches and *D* is the embedding dimension) are prepended with the CLS token:

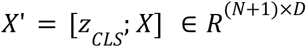

Before applying self-attention, the embeddings pass through a small transformation network consisting in a linear layer, ReLU activation and dropout:

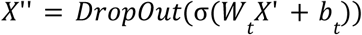

where *W_t_* ∈ *R*^*D*×*D*^ and *b_t_* ∈ *R^D^* are trainable parameters, and σ(·) represents the ReLU activation function. A LayerNorm operation is then applied through *X_norm_* = *LayerNorm*(*X*’’).

We then apply multi-head self-attention:

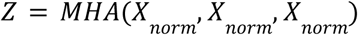

where MHA is described as

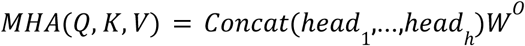

with each attention head computing

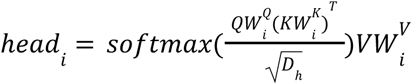

where 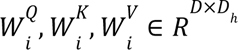 are learnable projection matrices, *D_h_* is the dimension per head and *Q*, *K*, *V* the query, key and value matrices.

Finally, we extract the representation as

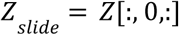

This final slide-level embedding is finally processed through a learnable linear head for our classification task as

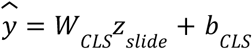

where *W_CLS_* ∈ *R*^*C*×*D*^ and *b_CLS_* ∈ *R^C^* are classification head parameters for *C* output classes.

As presented in this section, we extensively tested various combinations and aggregation strategies, including multiple instance learning approaches to learn representation directly at the slide-level such as DeepSets, gated attention and transformer-based attention. We also tried simpler yet robust permutation-invariant methods to aggregate image-level learned features. Our results (**Supplementary Table 1**) indicate that the simpler mean aggregation performed competitively, and in some cases, outperformed the more complex MIL-based models. A possible explanation for this observation is that MIL methods rely on learning instance-level statistics across patches, which may require a larger dataset to generalize effectively. Nonetheless, we hypothesize that MIL-based aggregation could offer advantages in larger-scale data configurations by even better capturing subtle, discriminative features patch-wise.

### Deep learning model hyperparameters and training

To ensure a robust and unbiased evaluation, we performed a stratified split of the slides in 4 subsets that maintained both class balance (*negative*, *submicroscopic*) and study balance (samples originated from two distinct studies: Asympto 1 and Asympto 2). This resulted in a 4-fold cross-validation setup with mutually exclusive test sets of 25% of the data with FoVs grouped by slides and no overlap between the four test subsets, thereby preventing any potential data leakage. For consistency, the same 4 folds were used to compare the combinations of encoders and aggregations, while trainings were performed 4 times with 4 random seeds. For optimization, the AdamW optimizer was used, with a weight decay of 0.01 to improve generalization. Various dropout probabilities were also tried across architectures, particularly in the learnable classification head, with the optimal value found to be 3 DropOut layers of respectively 0.5 probability for the first one, and 0.3 for the following 2. We evaluated different learning rate scheduling strategies, including cosine annealing and *ReduceLROnPlateau*. Still the best performances were obtained with a constant learning rate of 10 × 10^−5^. Data augmentation was applied to improve model generalization across experimental variations. A 512×512 fixed resized transformation, along with Random Rotation (0, 90, 120, 270 degrees to avoid black corners), Colour Jitter, Horizontal and Vertical Flip transforms were also applied. Furthermore, color jittering was employed to compensate for coloration shifts that might arise from experimental variations in staining and imaging conditions. To ensure efficiency when using pretrained models, we stored 10 randomly augmented images’ embeddings per image for the train set. For the test set, one embedding per image was stored, after resize & normalization. In our experiments, different transfer learning and fine-tuning strategies were evaluated. These included full fine-tuning and standard transfer learning (where only the classification head was trained). Given the limited amount of data available, transfer learning proved to be the most effective strategy, as full fine-tuning and unfreezing strategies led to overfitting. Furthermore, label smoothing was incorporated as an additional regularization technique due to the close phenotypic similarity between the *negative* and *submicroscopic* classes. By distributing a small portion of the label probability mass across classes, label smoothing encouraged the model to avoid overconfidence in its predictions. Formally, given a ground truth label *y* and the model’s predicted probability distribution *p*(*y*|*x*), label smoothing modifies the target distribution as follows:

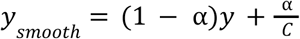

where *a* is the smoothing factor and *C* is the total number of classes. This adjustment helps prevent the model from becoming overly confident in its predictions and encourages it to learn more discriminative features between similar classes. To explore the best training configuration, we conducted an extensive grid search. The first stage of the search focused on hyperparameters such as learning rate, scheduler, minimum learning rate ETA, batch size, and two different architectures for the classification head - ranging from shallow to deeper MLP layers. Additionally, we evaluated different values of label smoothing, which played a crucial role in improving performance, particularly in handling class proximity. The search also included testing various backbone architectures. Once the best configuration was identified - one that yielded optimal performance while minimizing overfitting - we maintained all hyperparameters constant to systematically test the different aggregation strategies. This ensured a fair and deterministic comparison, with all models trained under similar conditions, including a constant learning rate of 10 × 10^−5^, no scheduling, a weight decay of 0. 01, and a batch size of 16. All reported experiments were conducted using NVIDIA A100-SXM4-80GB GPUs, ensuring computational efficiency and fast experimental iterating setup. Each training run was performed on a single GPU.

## Data Availability

All data produced in the present study are available upon reasonable request to the authors

## Data Availability

For reproducibility purposes, data will be made available on reasonable request.

## Code Availability

Codebase is available on Github under the following link: https://github.com/Saranga7/SMMALA

## Acknowledgements

This research has received support from ANR-10-IDEX-0001-02 PSL* Université Paris and from the Agence Nationale de la Recherche (ANR) under contracts ANR-23-CE19-0021-02 SMMALA and ANR-24-CE19-3949-01 COMPURES. This work was granted access to the HPC resources of IDRIS under the allocation 2020-AD011011495 made by GENCI.

## Author Contributions

NA, GC and AM performed blood smear sample collection, microscopy test and qPCR analyses. SKM and FL designed the smartphone application for sample acquisition. EL, NA and AA performed data acquisition. NB and SKM designed, coded, and performed all computational experiments and analyses. IS, FL and EC provided insights and help through the analyses. AG and GC conceived the project and supervised the work. NB and AG wrote the manuscript. All authors revised the manuscript.

## Declaration of generative AI and AI-assisted technologies in the manuscript preparation process

During the preparation of this work the author(s) sporadically used OpenAI ChatGPT on some parts of it in order to correct the English grammar. After using this tool/service, the author(s) reviewed and edited the content as needed and take(s) full responsibility for the content of the published article.

## References

Abba, Katharine, Jonathan J. Deeks, Piero Olliaro, et al. 2011. “Rapid Diagnostic Tests for Diagnosing Uncomplicated P. Falciparum Malaria in Endemic Countries.” The Cochrane Database of Systematic Reviews 2011 (7): CD008122.

Ando, D. Michael, Cory Y. McLean, and Marc Berndl. 2017. “Improving Phenotypic Measurements in High-Content Imaging Screens.” In bioRxiv. BioRxiv, July 10. 10.1101/161422.

Arevalo, John, Ellen Su, Robert van Dijk, Anne E. Carpenter, and Shantanu Singh. 2024. “Evaluating Batch Correction Methods for Image-Based Cell Profiling.” In bioRxiv. February 28. 10.1101/2023.09.15.558001.

Ashraf, Sania, Angie Kao, Cecilia Hugo, et al. 2012. “Developing Standards for Malaria Microscopy: External Competency Assessment for Malaria Microscopists in the Asia-Pacific.” Malaria Journal 11 (October): 352.

Bourou, Anis, Thomas Boyer, Kévin Daupin, et al. 2023. “PhenDiff: Revealing Subtle Phenotypes with Diffusion Models in Real Images.” In arXiv [eess.IV]. 10.48550/ARXIV.2312.08290.

Bourriez, Nicolas, Ihab Bendidi, Ethan Cohen, et al. 2023. “ChAda-ViT : Channel Adaptive Attention for Joint Representation Learning of Heterogeneous Microscopy Images.” arXiv [cs.CV], ahead of print. 10.48550/ARXIV.2311.15264.

Chandramohanadas, Rajesh, Yongkeun Park, Lena Lui, et al. 2011. “Biophysics of Malarial Parasite Exit from Infected Erythrocytes.” PloS One 6 (6): e20869.

Chaves, Alejandro Rojas, Yannelle Dossou, Armel Djènontin, et al. 2025. “Association between Asymptomatic Submicroscopic and Microscopic Malaria Infections and Anemia: A Study in Southern Benin.” PloS One 20 (1): e0317345.

Das, Dev Kumar, Madhumala Ghosh, Mallika Pal, Asok K. Maiti, and Chandan Chakraborty. 2013. “Machine Learning Approach for Automated Screening of Malaria Parasite Using Light Microscopic Images.” Micron (Oxford, England : 1993) 45 (February): 97–106.

Dekel, Elya, Dana Yaffe, Irit Rosenhek-Goldian, et al. 2021. “20S Proteasomes Secreted by the Malaria Parasite Promote Its Growth.” Nature Communications 12 (1): 1172.

Eijk, Anna Maria van, Kasia Stepniewska, Jenny Hill, et al. 2023. “Prevalence of and Risk Factors for Microscopic and Submicroscopic Malaria Infections in Pregnancy: A Systematic Review and Meta-Analysis.” The Lancet. Global Health 11 (7): e1061–e1074.

Esteva, Andre, Brett Kuprel, Roberto A. Novoa, et al. 2017. “Dermatologist-Level Classification of Skin Cancer with Deep Neural Networks.” Nature 542 (7639): 115–118.

Fillioux, Leo, Emilie Gontran, Jérôme Cartry, et al. 2023. “Spatio-Temporal Analysis of Patient-Derived Organoid Videos Using Deep Learning for the Prediction of Drug Efficacy.” In arXiv [cs.CV]. 10.48550/ARXIV.2308.14461.

Garcia, Lynne S. 2010. “Malaria.” Clinics in Laboratory Medicine 30 (1): 93–129.

Gilson, Paul R., Scott A. Chisholm, Brendan S. Crabb, and Tania F. de Koning-Ward. 2017. “Host Cell Remodelling in Malaria Parasites: A New Pool of Potential Drug Targets.” International Journal for Parasitology 47 (2-3): 119–127.

He, Kaiming, Xinlei Chen, Saining Xie, Yanghao Li, Piotr Dollár, and Ross Girshick. 2021. “Masked Autoencoders Are Scalable Vision Learners.” In arXiv [cs.CV]. 10.48550/ARXIV.2111.06377.

Hung, Jane, Stefanie C. P. Lopes, Odailton Amaral Nery, et al. 2017. “Applying Faster R-CNN for Object Detection on Malaria Images.” Conference on Computer Vision and Pattern Recognition Workshops. IEEE Computer Society Conference on Computer Vision and Pattern Recognition. Workshops 2017 (July): 808–813.

Ilse, Maximilian, Jakub M. Tomczak, and Max Welling. 2018. “Attention-Based Deep Multiple Instance Learning.” In arXiv [cs.LG]. 10.48550/ARXIV.1802.04712.

Kim, Hee E., Alejandro Cosa-Linan, Nandhini Santhanam, Mahboubeh Jannesari, Mate E. Maros, and Thomas Ganslandt. 2022. “Transfer Learning for Medical Image Classification: A Literature Review.” BMC Medical Imaging 22 (1): 69.

Lamiable, Alexis, Tiphaine Champetier, Francesco Leonardi, et al. 2023. “Revealing Invisible Cell Phenotypes with Conditional Generative Modeling.” Nature Communications 14 (1): 6386.

Lin Ouédraogo, André, Bronner P. Gonçalves, Awa Gnémé, et al. 2015. “Dynamics of the Human Infectious Reservoir for Malaria Determined by Mosquito Feeding Assays and Ultrasensitive Malaria Diagnosis in Burkina Faso.” The Journal of Infectious Diseases 213 (1): 90–99.

Litjens, Geert, Thijs Kooi, Babak Ehteshami Bejnordi, et al. 2017. “A Survey on Deep Learning in Medical Image Analysis.” Medical Image Analysis 42 (December): 60–88.

Lyu, Boyang, Thao Pham, Giles Blaney, et al. 2021. “Domain Adaptation for Robust Workload Level Alignment between Sessions and Subjects Using fNIRS.” Journal of Biomedical Optics 26 (2). 10.1117/1.JBO.26.2.022908.

Mathison, Blaine A., and Bobbi S. Pritt. 2017. “Update on Malaria Diagnostics and Test Utilization.” Journal of Clinical Microbiology 55 (7): 2009–2017.

McKinney, Scott Mayer, Marcin Sieniek, Varun Godbole, et al. 2020. “International Evaluation of an AI System for Breast Cancer Screening.” Nature 577 (7788): 89–94.

Moen, Erick, Dylan Bannon, Takamasa Kudo, William Graf, Markus Covert, and David Van Valen. 2019. “Deep Learning for Cellular Image Analysis.” Nature Methods 16 (12): 1233–1246.

Mosha, Jacklin F., Hugh J. W. Sturrock, Bryan Greenhouse, et al. 2013. “Epidemiology of Subpatent Plasmodium Falciparum Infection: Implications for Detection of Hotspots with Imperfect Diagnostics.” Malaria Journal 12 (July): 221.

Notomi, T., H. Okayama, H. Masubuchi, et al. 2000. “Loop-Mediated Isothermal Amplification of DNA.” Nucleic Acids Research 28 (12): E63.

Okell, Lucy C., Teun Bousema, Jamie T. Griffin, André Lin Ouédraogo, Azra C. Ghani, and Chris J. Drakeley. 2012. “Factors Determining the Occurrence of Submicroscopic Malaria Infections and Their Relevance for Control.” Nature Communications 3: 1237.

Paul, A., R. Pallavi, U. S. Tatu, and V. Natarajan. 2013. “The Bystander Effect in Optically Trapped Red Blood Cells due to Plasmodium Falciparum Infection.” Transactions of the Royal Society of Tropical Medicine and Hygiene 107 (4). 10.1093/trstmh/trt010.

Radford, Alec, Jong Wook Kim, Chris Hallacy, et al. 2021. “Learning Transferable Visual Models from Natural Language Supervision.” In arXiv [cs.CV]. 10.48550/ARXIV.2103.00020.

Raghu, Maithra, Chiyuan Zhang, Jon Kleinberg, and Samy Bengio. 2019. “Transfusion: Understanding Transfer Learning for Medical Imaging.” In arXiv [cs.CV]. 10.48550/ARXIV.1902.07208.

Rajaraman, Sivaramakrishnan, Sameer K. Antani, Mahdieh Poostchi, et al. 2018. “Pre-Trained Convolutional Neural Networks as Feature Extractors toward Improved Malaria Parasite Detection in Thin Blood Smear Images.” PeerJ 6 (April): e4568.

Scheeder, Christian, Florian Heigwer, and Michael Boutros. 2018. “Machine Learning and Image-Based Profiling in Drug Discovery.” Current Opinion in Systems Biology 10 (August): 43–52.

Scovino, Aline Miranda, Paulo Renato Rivas Totino, and Alexandre Morrot. 2022. “Eryptosis as a New Insight in Malaria Pathogenesis.” Frontiers in Immunology 13 (May): 855795.

Siméoni, Oriane, Huy V. Vo, Maximilian Seitzer, et al. 2025. “DINOv3.” In arXiv [cs.CV]. August 13. arXiv. http://arxiv.org/abs/2508.10104.

Sisquella, Xavier, Thomas Nebl, Jennifer K. Thompson, et al. 2017. “Ligand Binding to Erythrocytes Induce Alterations in Deformability Essential for Invasion.” eLife 6 (February). 10.7554/eLife.21083.

Suárez-Cortés, Pablo, Vikram Sharma, Lucia Bertuccini, et al. 2016. “Comparative Proteomics and Functional Analysis Reveal a Role of Plasmodium Falciparum Osmiophilic Bodies in Malaria Parasite Transmission.” Molecular & Cellular Proteomics : MCP 15 (10): 3243–3255.

Telfils, Rodeline, Akpéyédjé Yannelle Dossou, Armel Djènontin, et al. 2024. “Dynamics of Submicroscopic and Microscopic Asymptomatic Malaria Infection and Associated Factors: A Longitudinal Study in South Benin.” PloS One 19 (12): e0311217.

Torres, Katherine, Christine M. Bachman, Charles B. Delahunt, et al. 2018. “Automated Microscopy for Routine Malaria Diagnosis: A Field Comparison on Giemsa-Stained Blood Films in Peru.” Malaria Journal 17 (1): 339.

Vaswani, Ashish, Noam Shazeer, Niki Parmar, et al. 2017. “Attention Is All You Need.” In arXiv [cs.CL]. 10.48550/ARXIV.1706.03762.

Watson, Oliver J., Thu Nguyen-Anh Tran, Robert J. Zupko, et al. 2025. “Global Risk of Selection and Spread of Plasmodium Falciparum Histidine-Rich Protein 2 and 3 Gene Deletions.” Nature Medicine 31 (10): 3372–3379.

World Health Organization. 2010. Basic Malaria Microscopy: Tutor’s Guide. World Health Organization.

Wu, Lindsey, Lotus L. van den Hoogen, Hannah Slater, et al. 2015. “Comparison of Diagnostics for the Detection of Asymptomatic Plasmodium Falciparum Infections to Inform Control and Elimination Strategies.” Nature 528 (7580): S86–93.

Yang, Feng, Mahdieh Poostchi, Hang Yu, et al. 2020. “Deep Learning for Smartphone-Based Malaria Parasite Detection in Thick Blood Smears.” IEEE Journal of Biomedical and Health Informatics 24 (5): 1427–1438.

Yosinski, Jason, Jeff Clune, Yoshua Bengio, and Hod Lipson. 2014. “How Transferable Are Features in Deep Neural Networks?” arXiv [cs.LG], ahead of print. 10.48550/ARXIV.1411.1792.

Zaheer, Manzil, Satwik Kottur, Siamak Ravanbakhsh, Barnabas Poczos, Ruslan Salakhutdinov, and Alexander Smola. 2017. “Deep Sets.” In arXiv [cs.LG]. 10.48550/ARXIV.1703.06114.

Zhang, Yao, Changjin Huang, Sangtae Kim, et al. 2015. “Multiple Stiffening Effects of Nanoscale Knobs on Human Red Blood Cells Infected with Plasmodium Falciparum Malaria Parasite.” Proceedings of the National Academy of Sciences of the United States of America 112 (19): 6068–6073.

Zuccala, Elizabeth S., Timothy J. Satchwell, Fiona Angrisano, et al. 2016. “Quantitative Phospho-Proteomics Reveals the Plasmodium Merozoite Triggers Pre-Invasion Host Kinase Modification of the Red Cell Cytoskeleton.” Scientific Reports 6 (February): 19766.

