## Supplementary Material for "Detection of Malaria Infection from parasite-free blood smears"

### Supplementary Information

#### Supplementary Figures

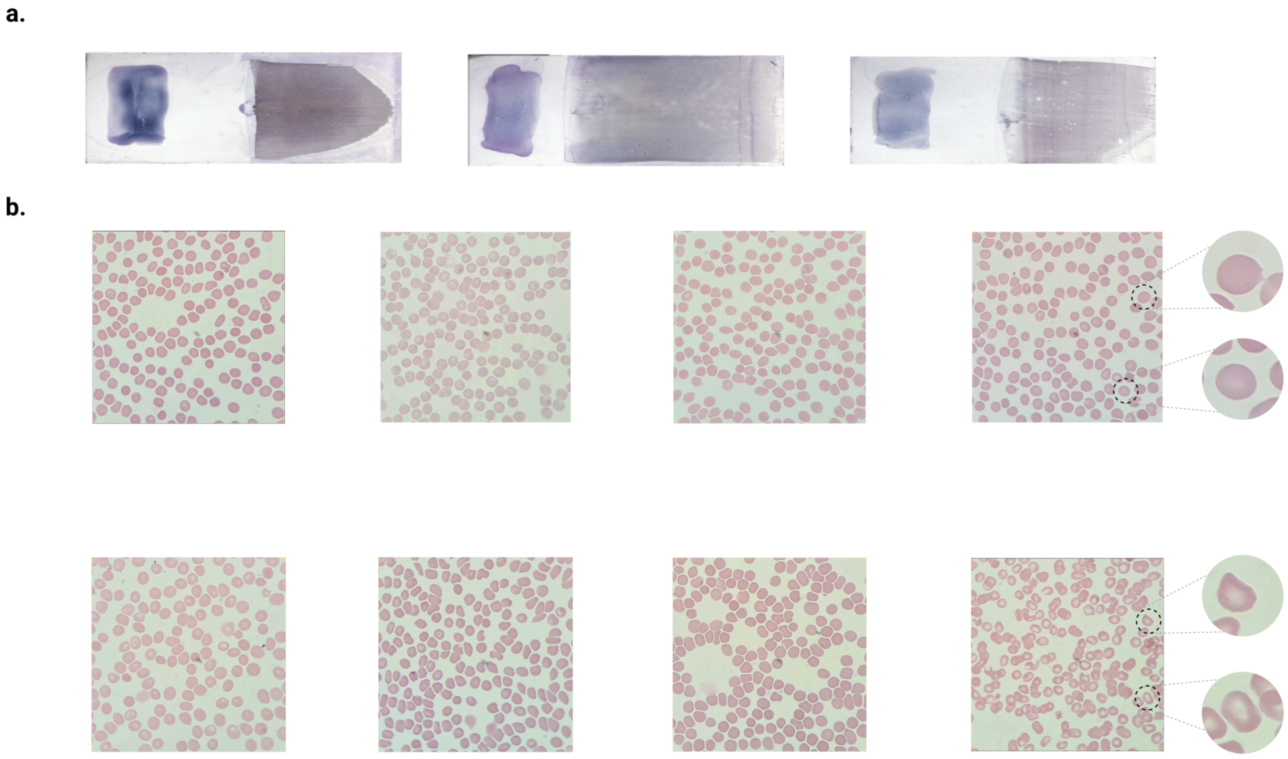

**Supplementary Figure 1: Representative raw thick and thin blood smear slides, together with high-magnification fields of view used for model training and evaluation a.** 3 examples of thick (left side of the slides) and thin (right side of the slides) blood smear slides stained with Giemsa **b.** Random fields-of-view (FoVs) acquired at ×1000 oil immersion magnification using a smartphone-mounted microscope for each diagnostic class: *submicroscopic* (bottom row), and *negative* (top row). *Submicroscopic* samples contain no visible parasites despite qPCR positivity, and *negative* samples are both microscopy and qPCR-negative. These images highlight the challenge of identifying submicroscopic infections through human inspection alone.

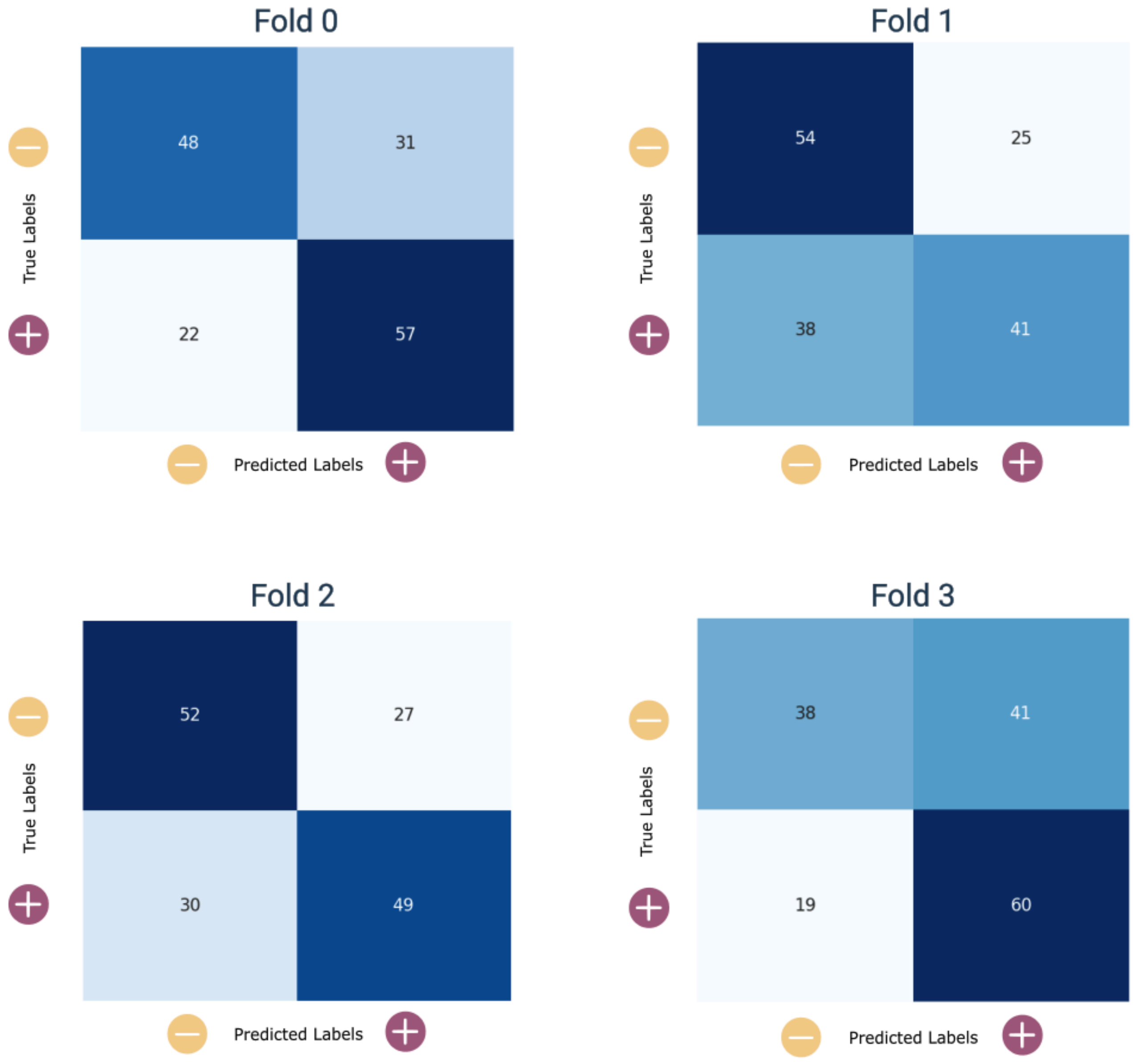


**Supplementary Figure 2: Confusion matrices for the best model** — selected based on accuracy and sensitivity — across 4-fold cross-validation. Matrices are shown per fold, each evaluated with a distinct random seed.

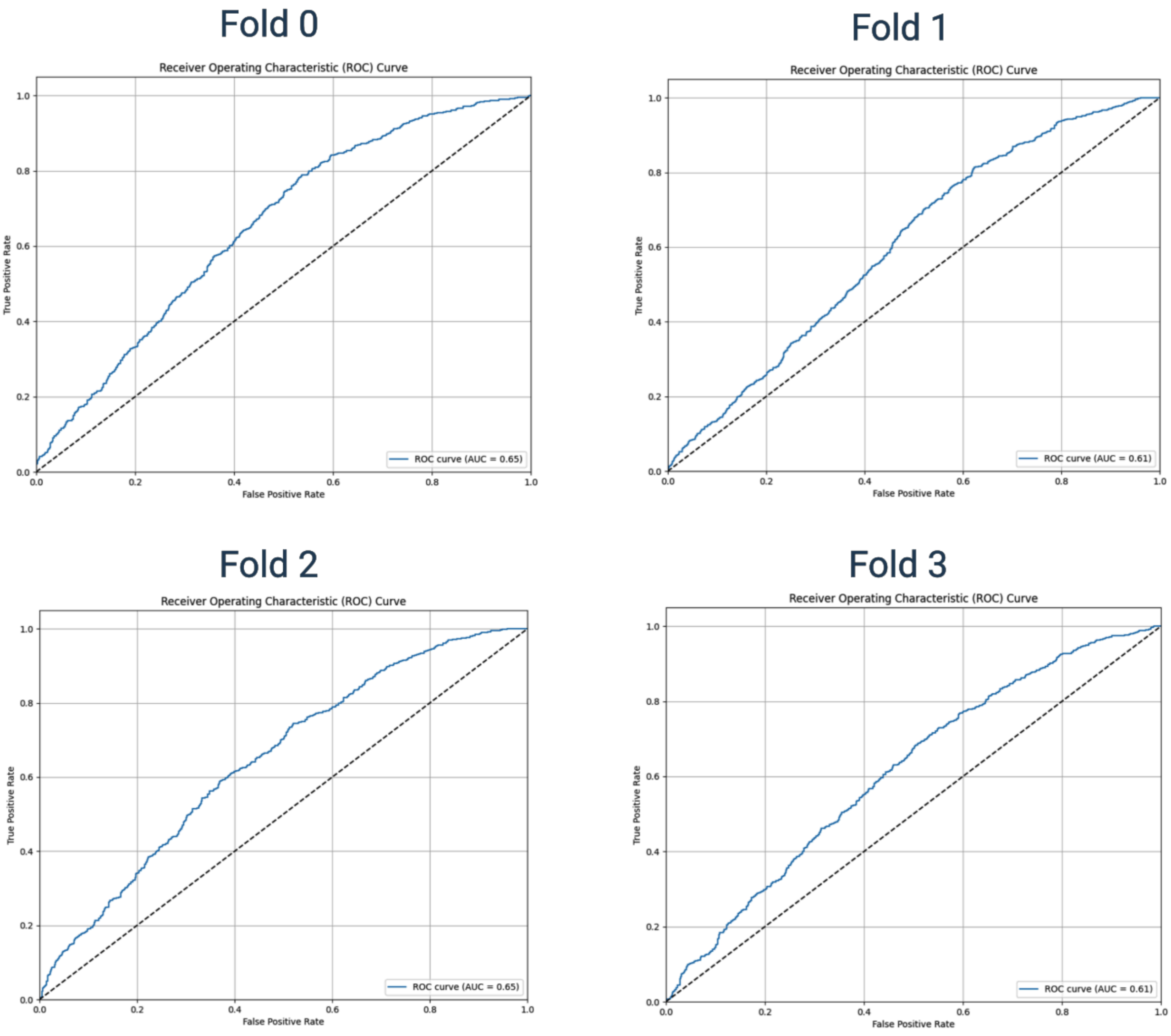


**Supplementary Figure 3: Receiver Operating Curves for the best model** — selected based on accuracy and sensitivity — across 4-fold cross-validation. Curves are shown per fold, each evaluated with a distinct random seed.

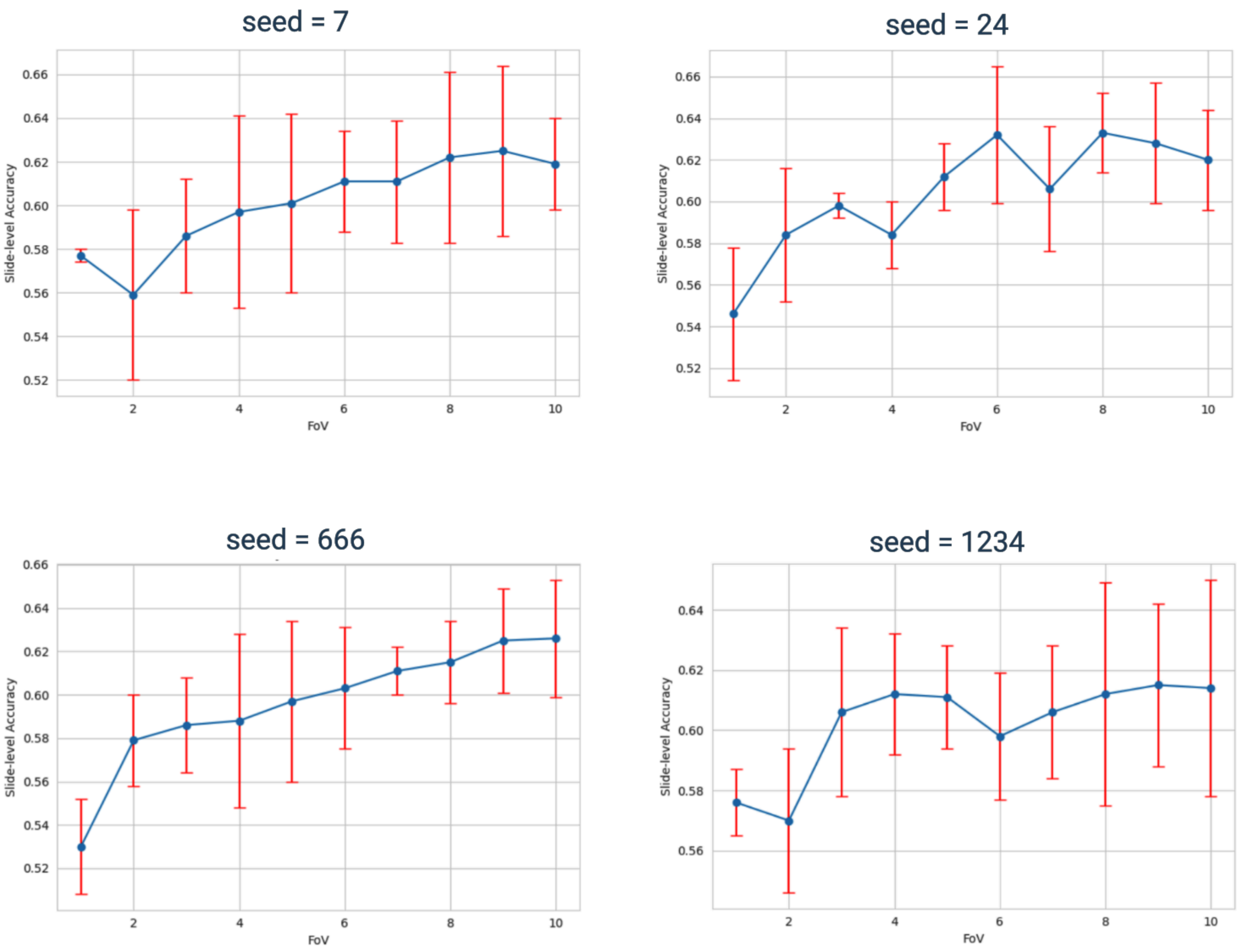


**Supplementary Figure 4: Accuracy on FoV number curves for the best model** — selected based on accuracy and sensitivity — across 4-fold cross-validation and 4 different seeds. Curves are shown per seed, each evaluated on the 4 folds.

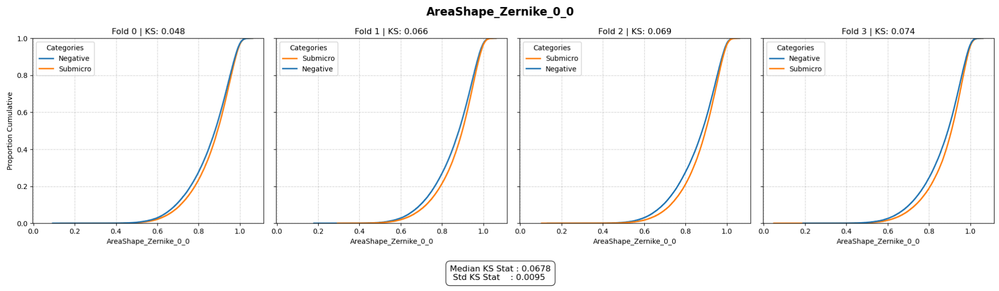

**CentralPallorRatio**

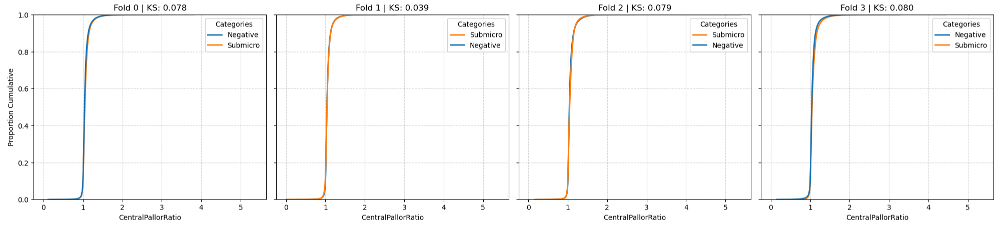


Median KS Stat : 0.0787
Std KS Stat : 0.0176

**AreaShape_HuMoment_0**

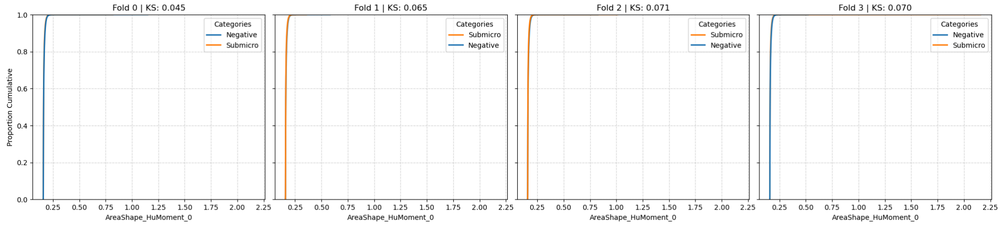


Median KS Stat : 0.0677
Std KS Stat : 0.0104

**AreaShape_Zernike_2_0**

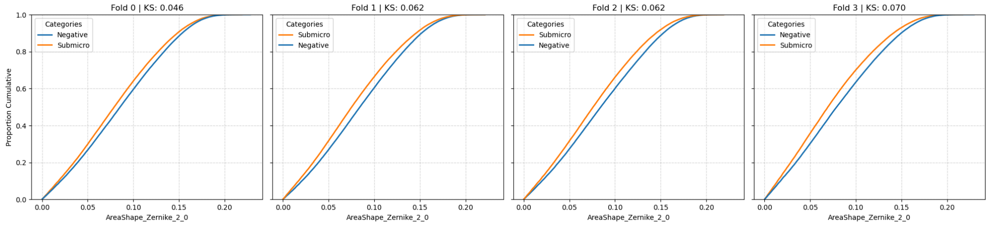


Median KS Stat : 0.0620
Std KS Stat : 0.0089

**AreaShape_HuMoment_3**

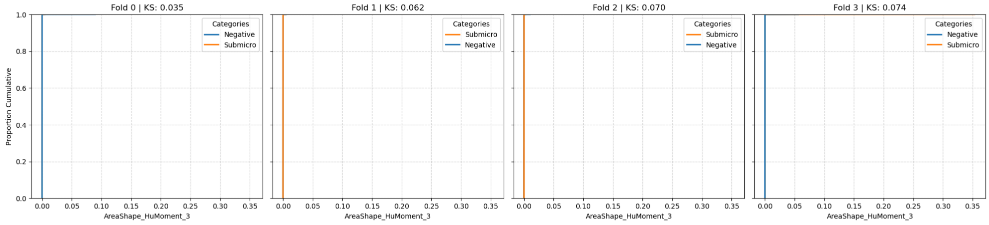


Median KS Stat : 0.0662
Std KS Stat : 0.0152

**AreaShape_Compactness**

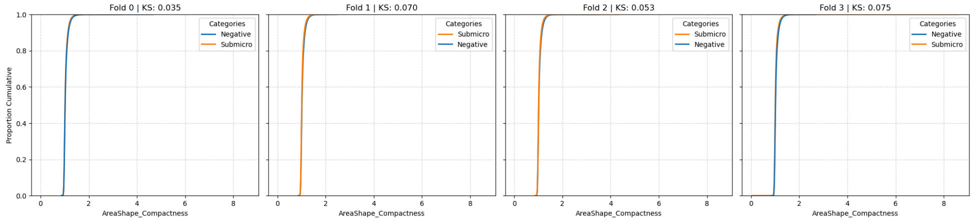


Median KS Stat : 0.0618
Std KS Stat : 0.0159

**AreaShape_FormFactor**

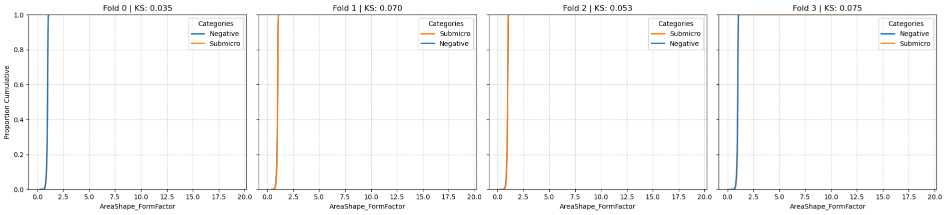


Median KS Stat : 0.0618
Std KS Stat : 0.0159

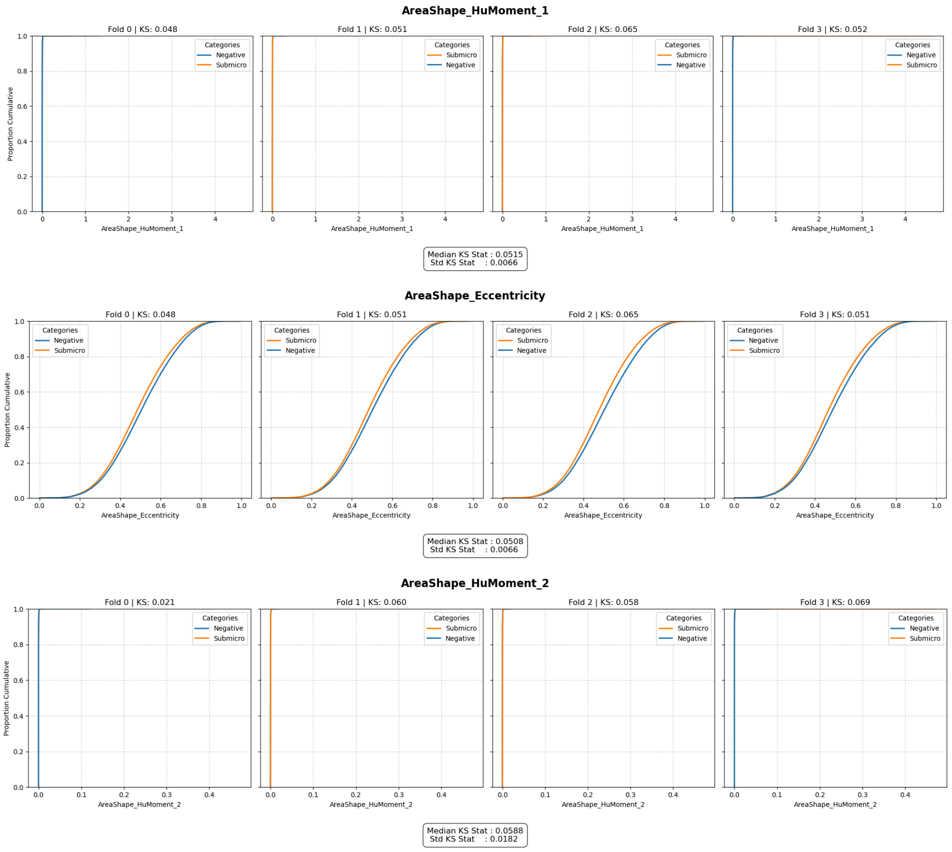


**Supplementary Figure 5: Empirical Cumulative Distribution Functions for the top 10 features with highest KS statistic.** ECDF plots with related KS stat & p-value are computed per fold on true data samples.

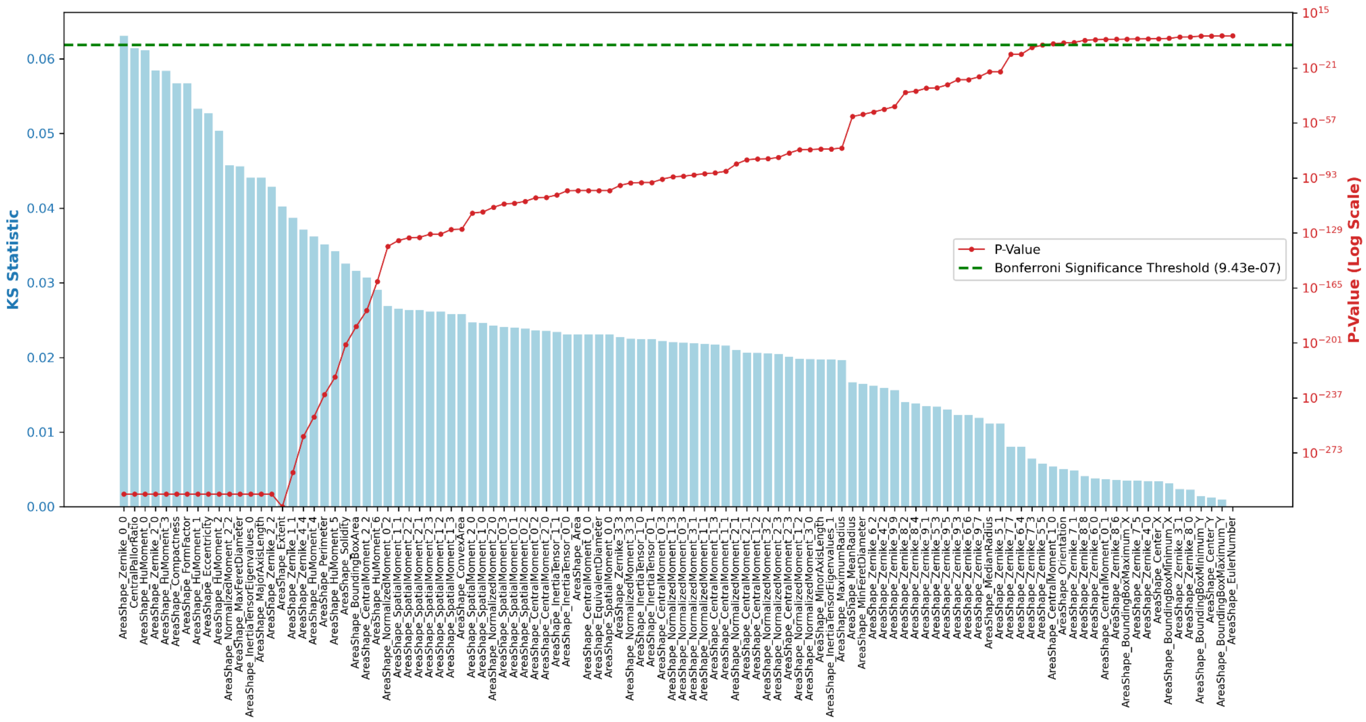


**Supplementary Figure 6: Statistical ranking of morphological features distinguishing true samples.** Features are ranked by their Kolmogorov–Smirnov (KS) statistic (blue bars, left axis), representing the magnitude of the difference between negative and submicroscopic sample distributions. The corresponding p-values (red line, right axis; logarithmic scale) indicate statistical significance. The horizontal dashed green line denotes the Bonferroni-corrected significance threshold ($P < 9.43 \times 10^{-7}$), adjusted for multiple hypothesis testing across $n=106$ features. Features with p-values above this threshold (above the green line) are automatically rejected.

#### Supplementary Tables

| Model | Slide Aggregation | Accuracy | F1 | Specificity | Sensitivity |
|---|---|---|---|---|---|
| Custom CNN (small) | mean | 0.501 ± 0.012 | 0.345 ± 0.032 | 0.567 ± 0.494 | 0.434 ± 0.503 |
| Custom CNN (small) | median | 0.501 ± 0.014 | 0.345 ± 0.033 | 0.566 ± 0.493 | 0.435 ± 0.503 |
| Custom CNN (small) | vote | 0.500 ± 0.011 | 0.345 ± 0.034 | 0.568 ± 0.492 | 0.432 ± 0.500 |
| Custom CNN (big) | mean | 0.500 ± 0.000 | 0.333 ± 0.000 | 1.000 ± 0.000 | 0.000 ± 0.000 |
| Custom CNN (big) | median | 0.500 ± 0.000 | 0.333 ± 0.000 | 1.000 ± 0.000 | 0.000 ± 0.000 |
| Custom CNN (big) | vote | 0.500 ± 0.000 | 0.333 ± 0.000 | 1.000 ± 0.000 | 0.000 ± 0.000 |
| ResNet50 | mean | 0.556 ± 0.036 | 0.534 ± 0.038 | 0.350 ± 0.070 | 0.763 ± 0.082 |
| ResNet50 | median | 0.564 ± 0.031 | 0.543 ± 0.034 | 0.354 ± 0.064 | 0.775 ± 0.072 |
| ResNet50 | vote | 0.559 ± 0.033 | 0.538 ± 0.034 | 0.361 ± 0.067 | 0.756 ± 0.081 |
| DINOv3 ViT-H/16+ | mean | 0.579 ± 0.043 | 0.569 ± 0.048 | 0.441 ± 0.076 | 0.717 ± 0.057 |
| DINOv3 ViT-H/16+ | median | 0.577 ± 0.045 | 0.566 ± 0.052 | 0.429 ± 0.089 | 0.725 ± 0.056 |
| DINOv3 ViT-H/16+ | vote | 0.587 ± 0.040 | 0.579 ± 0.044 | 0.465 ± 0.081 | 0.710 ± 0.063 |
| DINOv3 ViT-S/16 | mean | 0.593 ± 0.028 | 0.590 ± 0.028 | 0.552 ± 0.068 | 0.634 ± 0.102 |
| DINOv3 ViT-S/16 | median | 0.600 ± 0.035 | 0.595 ± 0.036 | 0.527 ± 0.081 | 0.672 ± 0.100 |
| DINOv3 ViT-S/16 | vote | 0.598 ± 0.034 | 0.594 ± 0.035 | 0.571 ± 0.084 | 0.624 ± 0.106 |
| DINOv3 ViT-S/16+ | mean | 0.602 ± 0.032 | 0.599 ± 0.033 | 0.520 ± 0.047 | 0.685 ± 0.035 |
| DINOv3 ViT-S/16+ | median | 0.589 ± 0.039 | 0.585 ± 0.039 | 0.500 ± 0.043 | 0.678 ± 0.054 |
| DINOv3 ViT-S/16+ | vote | 0.587 ± 0.041 | 0.585 ± 0.041 | 0.528 ± 0.044 | 0.647 ± 0.068 |
| **DINOv3 ViT-B/16** | **mean** | **0.620 ± 0.025** | 0.616 ± 0.025 | 0.567 ± 0.070 | 0.672 ± 0.096 |
| DINOv3 ViT-B/16 | median | 0.609 ± 0.039 | 0.605 ± 0.038 | 0.539 ± 0.062 | 0.680 ± 0.114 |
| DINOv3 ViT-B/16 | vote | 0.611 ± 0.030 | 0.607 ± 0.031 | 0.581 ± 0.071 | 0.640 ± 0.121 |
| DINOv3 ViT-B/16 | deepset | 0.587 ± 0.013 | 0.583 ± 0.014 | 0.507 ± 0.069 | 0.668 ± 0.074 |
| DINOv3 ViT-B/16 | gated_attention | 0.595 ± 0.012 | 0.593 ± 0.012 | 0.547 ± 0.057 | 0.643 ± 0.068 |
| DINOv3 ViT-B/16 | top-$k$ | 0.584 ± 0.015 | 0.577 ± 0.019 | 0.478 ± 0.080 | 0.691 ± 0.074 |
| DINOv3 ViT-B/16 | transformer_attention | 0.594 ± 0.027 | 0.590 ± 0.026 | 0.536 ± 0.071 | 0.652 ± 0.093 |

**Supplementary Table 1:. Performance metrics for models using *simple* and complex (last four rows) slide-level aggregation.** Reported accuracy corresponds to the overall mean across 4 stratified folds of the per-fold mean accuracy over 4 runs with distinct random seeds. Values are presented as mean ± std.
